# Comparison of common respiratory pathogen detection in nasopharyngeal swabs, saline gargle specimens, pleural effusion and lower respiratory tract samples

**DOI:** 10.1101/2022.10.24.22281433

**Authors:** Olympia E. Anastasiou

## Abstract

Multiplex PCR panels have been used for the diagnosis of viral respiratory infections in the last years. While the types of manufacturer validated and thus officially approved materials are usually limited, the tested materials in the clinical routine or studies often vary, which presents a challenge in light of the new EU-IVDR guideline. Aim of our present study was to evaluate if testing of lower respiratory tract (LRT) or saline gargle specimens (SGS) provided an advantage compared to the testing of nasopharyngeal swabs (NPS) and if the testing of pleural effusions (PE) provided any advantage compared to the testing of LRT samples.

We included 367 NPS vs LRT cases, of which 202 (55%) were negative in both samples, 108 (29%) were positive in both samples, 28 (8%) had a positive NPS and a negative LRT and 29 (8%) had a negative NPS but a positive LRT, with no significant differences between immunocompetent and immunosuppressed cases. We included 46 NPS vs SGS cases, of which 18 (39%) were negative in both samples, 18 (39%) were positive in both samples, 4 (9%) had a positive NPS and a negative SGS and 6 (13%) had a negative NPS but a positive SGS. Out of the 82 tested PE samples, only one (1%) was positive for Influenza B RNA (detected in the PE but not LTR), while for 5 positive LTR samples no viral genome could be detected in the PE. The samples were tested with the FTD respiratory viral panel for common respiratory viruses.

Testing of a lower respiratory tract sample after a negative upper respiratory tract sample may have an incremental diagnostic value. Gargle and nasopharyngeal swab samples seem to have a comparable diagnostic performance, while pleural effusion is a substandard material for the diagnosis of common respiratory virus infections.

## Introduction

Respiratory tract infections have always been and continue to be a significant cause of overall morbidity and mortality [1]. In recent years, our diagnostic capabilities have been enhanced including multiplex PCR, enabling the testing of multiple potential pathogens. To date several clinical studies have used a plethora of different sample types from the upper and lower respiratory tract to pinpoint the agents causing the respiratory infection, a universally accepted golden standard does not exist [2].

The multiplex PCR tests for all their advantages have also drawbacks such as the high cost of implementation and the need for specialized laboratories and personnel. Furthermore the validation of these assays through the manufacturer is restricted due to practical reasons (e.g. potential revenue vs expenses) to certain matrices. For respiratory panels, the validated material often is the nasopharyngeal swab [3–5] but may also include other materials such as bronchoalveolar lavage (BAL)[6]. Taking into account the implementation of the new European Union In-Vitro Diagnostic Device Regulation (IVDR), analyzing other than the manufacturer approved sample matrices will require a laborious, time-consuming and expensive in-house validation, since the test combination with the non-approved type of sample is perceived as a laboratory developed test [7]. The costs and time invested for such a validation in the case a multiplex PCR is prohibitive for most laboratories. Thus, choosing the right platform and the right materials for testing is very important.

Aim of our present study was to evaluate if the testing of lower respiratory tract (LRT) or saline gargle specimens (SGS) provided an advantage compared to the testing of nasopharyngeal swabs (NPS) and if the testing of pleural effusions (PE) provided any advantage compared to the testing of LRT samples.

## Patients and Methods

We performed a retrospective chart review on 367 cases of 305 patients, where a NPS was taken within a week of an LRT, 46 cases of 46 patients, where an NPS was taken within a week of a SGS and 31 cases of 31 patients, where a PE was taken within a week of a LRT sample. The 31 PE cases were part of an initial group of 82 tested PE samples. The SGS was produced after gargling with 10 ml of a NaCl 0.9% solution. For the purposes of the analysis LRT samples included both BAL and endotracheal aspirates (EA), as we had previously shown that their diagnostic performance is comparable [8]. The above-mentioned cases were identified from patient samples, which were tested in the Institute of Virology of University Hospital Essen, Germany, from June 2013 to June 2022 with the FTD respiratory viral panel (fast track diagnostics [FTD]; Siemens) for common respiratory viruses, which includes internal controls. The respiratory panel included testing for adenoviruses, coronaviruses (HCoV-OC43, HCoV-NL63, HCoV-229E, and HCoV-HKU1), enteroviruses, human metapneumovirus (HMPV), bocavirus, influenza viruses (A/H1N1/B), parainfluenza viruses (type 1–4), rhinoviruses and respiratory-syncytial virus (RSV). The differentiation between enteroviruses and rhinoviruses, when necessary, was performed using the Virella Entero LC 2.0 PCR assay (Gerbion). Nucleid acid extraction was performed using MagNA pure (Roche). Demographic and clinical data were taken from patient charts. Statistical analysis was performed using SPSS software (v23, SPSS Inc., Chicago, IL, USA) and the platform VassarStats (http://vassarstats.net). Two-tailed p values less than 0.05 were considered to be statistically significant. This retrospective study was carried out in accordance with the Declaration of Helsinki and the guidelines of the International Conference for Harmonization for Good Clinical Practice and was approved by the ethics and research committee of the University Hospital of Essen-Duisburg (BO 20-9265-BO, 14.04.2020).

## Results

### In 13% of cases, a respiratory virus could be detected in an LRT but not in the corresponding NPS: no significant difference between immunocompetent vs immunosuppressed cohorts

In the comparison between NPS and LRT, we included 367 cases, of which 202 (55%) were negative in both samples, 108 (29%) were positive in both samples, 28 (8%) had a positive NPS and a negative LRT and 29 (8%) had a negative NPS but a positive LRT (Figure 1A). The measure of agreement, expressed as Kappa (k), was 0.668. A 60% (n=219) of the cases belonged to male patients and 40% (n=148) to female, while 38% (140) of the cases belonged to immunocompetent patients and 62% (n=227) to immunocompromised, of which 70% (n=159) to lung transplant recipients. Data on the 29 discrepant cases can be found in Supplement (Table S1).

**Figure 1:**
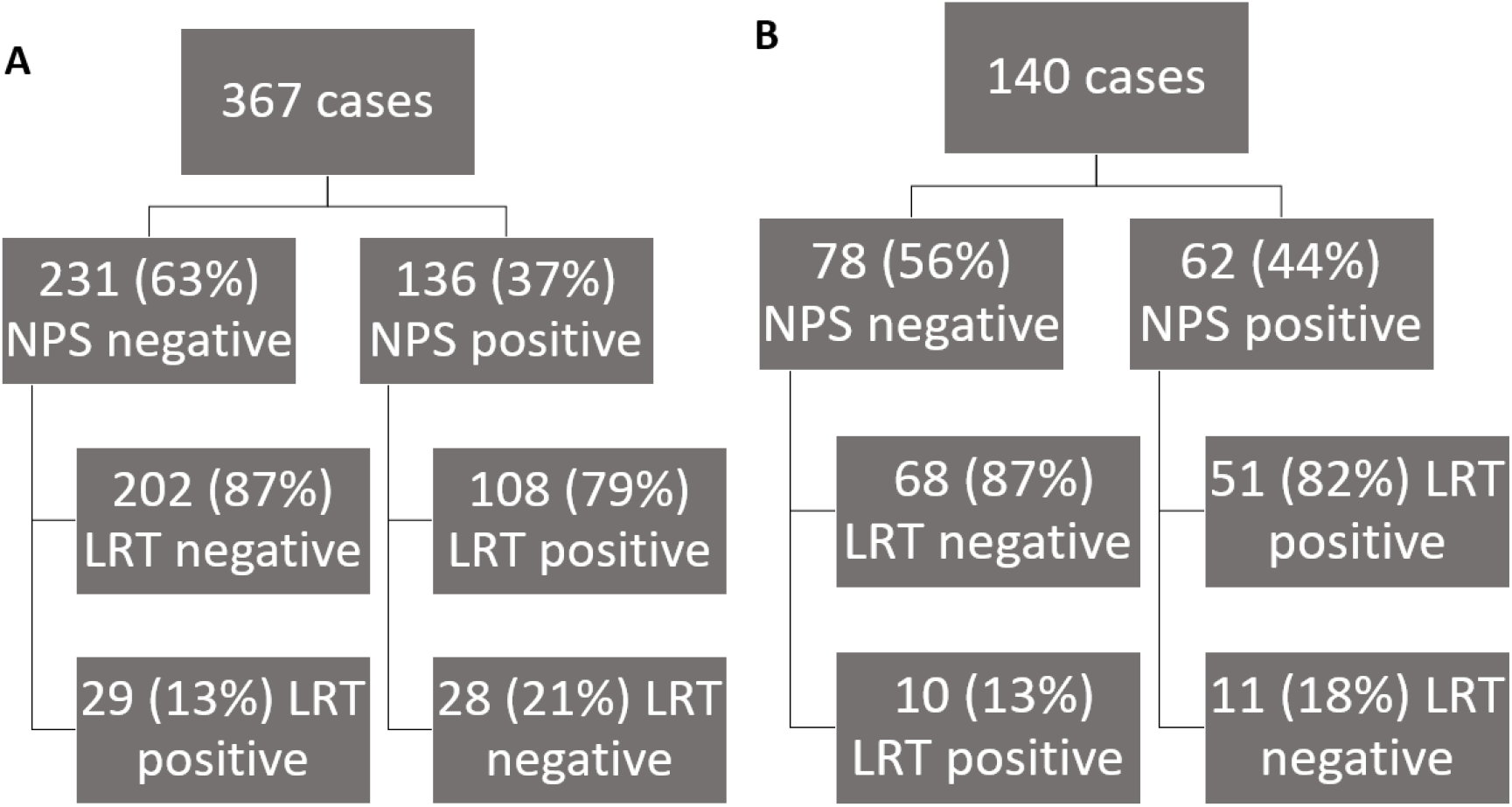
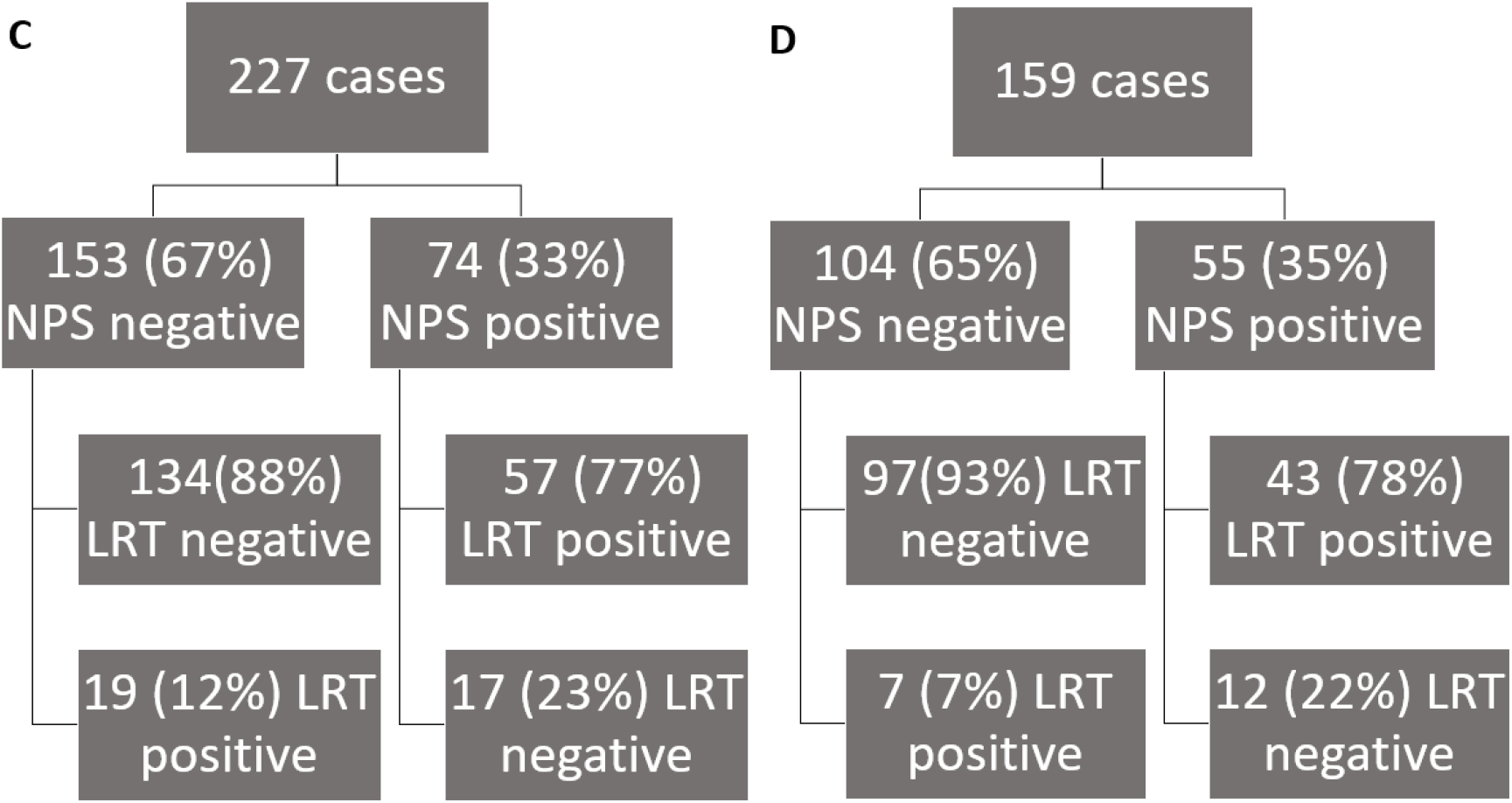
Distribution of positive and negative results for common respiratory viruses in nasopharyngeal swab samples (NPS) and lower respiratory tract samples (LRT) in all cases (A), in cases from immunocompetent hosts (B), in cases from immunosuppressed hosts (C) and in cases from lung transplant recipients (D).

After stratifying the cases according to the immune status of the patients involved, we found the following. Of the 140 cases included in the immunocompetent group, 68 (49%) were negative in both samples, 51 (36%) were positive in both samples, 11 (8%) had a positive NPS and a negative LRT and 10 (8%) had a negative NPS but a positive LRT (Figure 1B). Of the 227 cases included in the immunosuppressed group, 134 (59%) were negative in both samples, 57 (25%) were positive in both samples, 17 (8%) had a positive NPS and a negative LRT and 19 (8%) had a negative NPS but a positive LRT (Figure 1C). When focusing only on the lung transplant recipients, of the 159 cases, 97 (61%) were negative in both samples, 43 (27%) were positive in both samples, 12 (8%) had a positive NPS and a negative LRT and 7 (4%) had a negative NPS but a positive LRT (Figure 1D). The measurements of agreement as expressed by kappa were 0.696 for the immunocompetent cohort, 0.642 for the immunosuppressed cohort and 0.730 for the lung transplant recipients.

In NPS the most commonly detected viruses (only or dominant virus in cases where multiple viruses were detected) were rhinoviruses (n=53, 39%), followed by influenza virus A and coronavirus OC43 (each n=16, 12%), followed by RSV (n=11, 8%). In LPR the most commonly detected viruses (only or dominant virus in cases where multiple viruses were detected) were also rhinoviruses (n=45, 33%), followed by influenza virus A (n=15, 11%), followed coronavirus OC43 and adenoviruses (each n=12, 9%). Multiple viruses were detected in 4% (n=16) of the NPS and 6% (n=22) of the LPR. In cases were both samples were positive the Ct values for the detected virus or for the virus with the lowest Ct value in cases, where multiple viruses were detected, were lower in the LRT compared to the NPS [25 (21-30) vs 27 (23-32), p=0.008].

### Almost a third of cases in children with negative NPS samples are positive for common respiratory viruses in the corresponding LRT

After dividing the cases according to age (children vs adults), we found the following. Of the 251 cases included in the adult group, 156 (62%) were negative in both samples, 63 (25%) were positive in both samples, 20 (8%) had a positive NPS and a negative LRT and 12 (5%) had a negative NPS but a positive LRT (Figure 2A). Of the 116 cases included in the children group, 46 (40%) were negative in both samples, 45 (39%) were positive in both samples, 8 (7%) had a positive NPS and a negative LRT and 17 (15%) had a negative NPS but a positive LRT (Figure 2B). The measurements of agreement as expressed by kappa were 0.705 for the adult cohort and 0.572 for the children.

**Figure 1:**
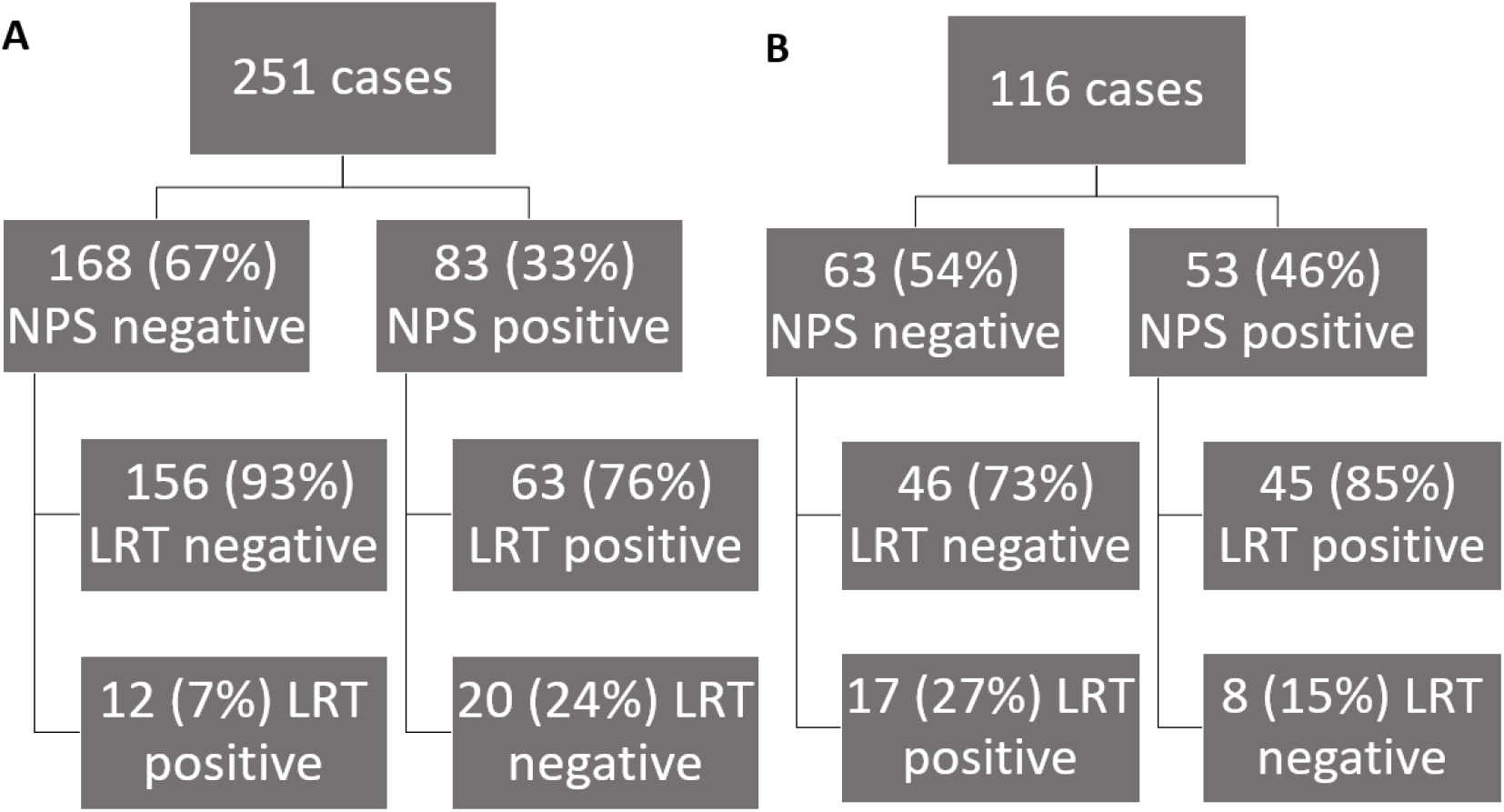
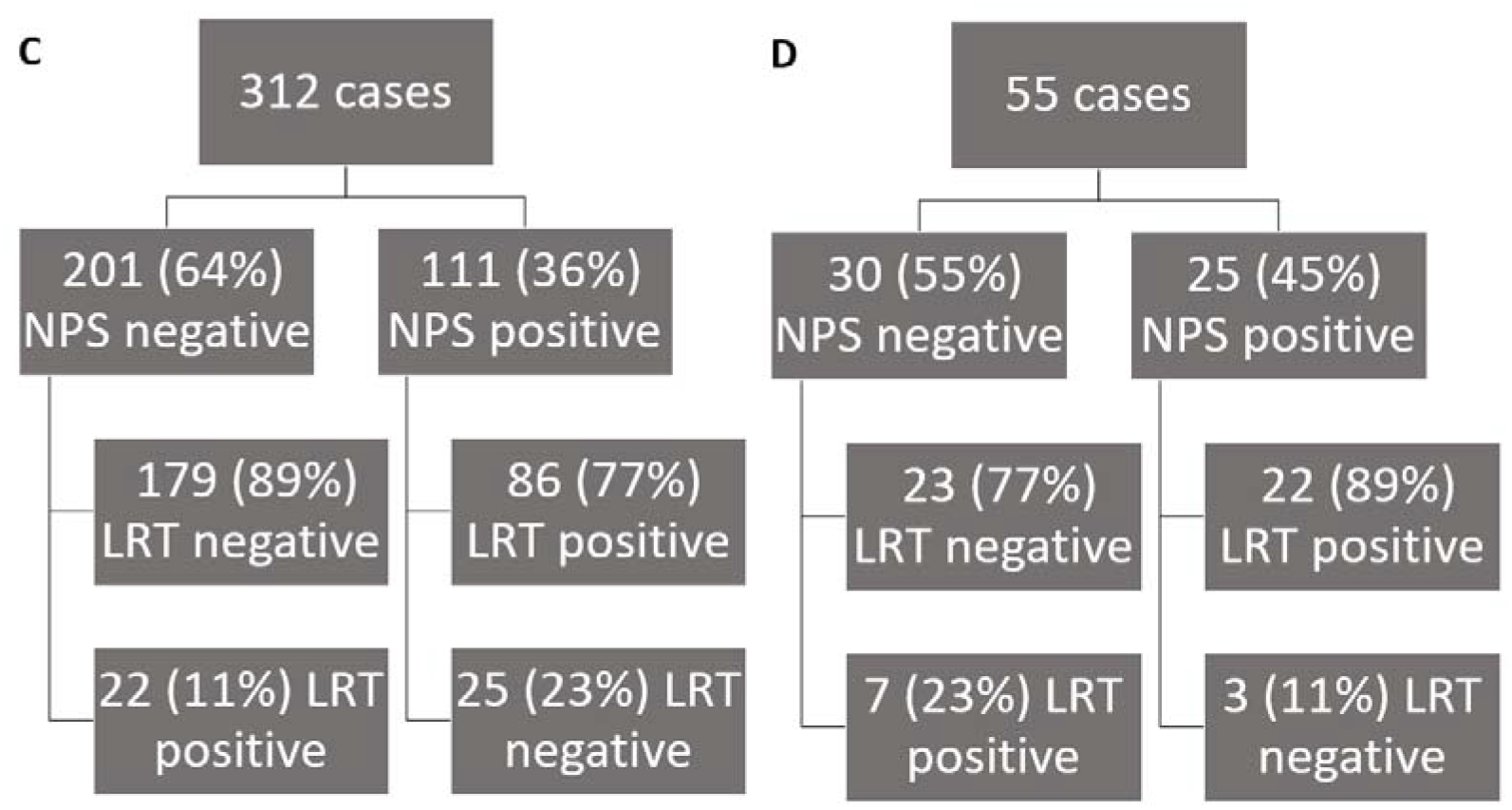
Distribution of positive and negative results for common respiratory viruses in nasopharyngeal swab samples (NPS) and lower respiratory tract samples (LRT) in cases of adult patients (A), in cases of children (B), in cases where bronchoalveolar lavage (BAL) was evaluated (C) and in cases where endotracheal aspirates (EA) were evaluated (D).

Furthermore, we stratified our cohort in cases with a BAL vs EA sample. Of the 312 cases included in the BAL group, 179 (57%) were negative in both BAL and NPS, 86 (28%) were positive in both samples, 22 (7%) had a positive NPS and a negative BAL and 25 (8%) had a negative NPS but a positive BAL (Figure 2C). Of the 55 cases included in the EA group, 23 (42%) were negative in both samples, 22 (40%) were positive in both samples, 7 (13%) had a positive NPS and a negative EA and 3 (6%) had a negative NPS but a positive EA (Figure 2D). The measurements of agreement as expressed by kappa were 0.669 for the BAL cohort and 0.638 for the EA cohort.

### NPS and SGS have a comparable diagnostic performance for common respiratory viruses

Next we evaluated, if SGS had a comparable diagnostic performance for common respiratory viruses to NPS. In the comparison between NPS and SGS, we included 46 cases, of which 18 (39%) were negative in both samples, 18 (39%) were positive in both samples, 4 (9%) had a positive NPS and a negative SGS and 6 (13%) had a negative NPS but a positive SGS. The measure of agreement, expressed as Kappa, was 0.566. A 54% (n=25) of the patients were male and 46% (n=21). Data on the 10 discrepant cases can be found in Supplement (Table S2).

### Pleura effusion is a substandard material for the diagnosis of lower respiratory infection through common respiratory viruses

Out of the 82 tested PE samples, only one (1%) was positive for a common respiratory virus. Influenza B RNA with a Ct value of 35 could be detected in the PE but not the subsequently tested BAL. In the comparison between PE and LTR samples, we included 31 cases, of which 25 (81%) were negative in both samples, 0 (0%) were positive in both samples, 1 (3%) had a positive PE and a negative LRT and 5 (16%) had a negative PR but a positive LPT. The measure of agreement, expressed as Kappa, was −0.057. A 52% (n=16) of the patients were male and 48% (n=15) were female. Data on the 6 discrepant cases can be found in Supplement (Table S3).

## Discussion

When comparing paired upper and lower respiratory tract samples, in 13% of cases, a respiratory virus could be detected in the lower but not in the upper respiratory tract. We observed no significant difference between immunocompetent (13%) vs immunosuppressed (12%) cases, while such a discrepancy was rarer (7%) in lung transplant recipients. Additionally in 62% and 34% of the above mentioned discrepant paired samples the Ct value of the detected viral pathogen was high (>30) or very high (>35) respectively, thus corresponding to a low or very low viral load, the clinical significance of which is unclear. Our results indicate a high degree of accordance between lower and upper respiratory tract samples concerning common respiratory viral infections. This is largely in accordance with previous studies focusing on immunocompromised and immunocompetent hosts [9, 10], on patients with COVID-19 induced respiratory failure[11] or suspected SARS-CoV-2 infection [12]. In general, testing a lower respiratory sample after obtaining a positive upper respiratory sample is superfluous for the purpose of diagnosing a viral respiratory infection in most clinical settings. However, a case can be made for testing a lower respiratory sample in the presence of a negative upper respiratory sample, if there is a substantiated clinical suspicion for a lower respiratory disease.

When comparing paired nasopharyngeal swabs and saline gargle specimens, we found that both sample types have a comparable diagnostic performance for common respiratory viruses. This is in accordance with the results of a previous study using an transcription-reverse transcription concerted reaction influenza assay [13], while a study comparing saliva samples and gargle samples showed that the first outperformed the latter compared to nasopharyngeal swabs in detecting SARS-CoV-2 RNA [14]. Overall, gargle samples seem to be an alternative to nasopharyngeal swabs, if they are already included in the manufacturer validated materials for an assay. However, since their diagnostic performance is comparable to those of nasopharyngeal swabs, an in-house validation seems to be a waste of resources.

Our results indicate that pleura effusion is a substandard material for the diagnosis of lower respiratory infection through common respiratory viruses. Out of the 82 tested samples, only one (1%) was positive for Influenza B RNA with a very low viral load (Ct=35). The virus was detected in the pleural effusion but not in the subsequently performed BAL. Interestingly in the case of 5 positive lower respiratory tract samples with Ct values ranging from 16 to 31, no viral genome could be detected in the PE. The non-existent yield of respiratory virus positive pleural effusion samples has been previously indicated in two studies presented in conferences, one looking at 138 patients with pleural effusions, which found no significant respiratory viral titres in the effusion [15] and another study focusing on pleural effusion after post coronary artery bypass grafting, which also did not find any DNA or RNA of common respiratory viruses in 48 samples [16]. Thus, validating pleural effusion as a material for common respiratory viruses assays and subsequently routinely testing the material is counterproductive.

The study has limitations due to its restrospective design. The paired samples were not obtained simultaneously. However, the brief interval (median 1 day with interquartile range of 0 to 3 days) between sample collections makes this limitation unlikely to have a significant impact on our findings. The number of paired samples in the cases of NPS vs SGS and PE vs LTR is small; as such, our results should be validated by future studies. Furthermore, our patient cohort is comprised by a disproportionate number of immunosuppressed individuals and especially lung transplant recipients, both due to the focus of our hospital but also the type of patient with a high likelihood of undergoing multiple testings for common respiratory viruses.

In conclusion, testing of a lower respiratory tract sample after a negative respiratory tract sample may have an incremental diagnostic value. Gargle and nasopharyngeal swab samples seem to have a comparable diagnostic performance, while pleural effusion is a substandard material for the diagnosis of common respiratory virus infections.

## Data Availability

Aggregated data produced in the present study are available upon reasonable request to the authors

## References

1. Estimates, G. H., Deaths by cause, age, sex, by country and by region, 2000-2016. 2018.

2. Hou, N.; Wang, K.; Zhang, H.; Bai, M.; Chen, H.; Song, W.; Jia, F.; Zhang, Y.; Han, S.; Xie, B., Comparison of detection rate of 16 sampling methods for respiratory viruses: a Bayesian network meta-analysis of clinical data and systematic review. BMJ Glob Health 2020, 5, (11).

3. Biomerieux, Syndromic Testing from Biofire: The Right Test, The First Time. In 2022.

4. Pathofinder, RespiFinder 2SMART: Including detection of SARS-CoV-2 and MERS-CoV. In 2022.

5. Diagnostics, F. T., FTD Pespiratory Pathogens 21: Beipackzettel. In 2022.

6. Luminex, Luminex NxTAG Assays: Molekulare Multiplex-PCR von Infektionserregern. In 2022.

7. Cobbaert, C.; Capoluongo, E. D.; Vanstapel, F. J. L. A.; Bossuyt, P. M. M.; Bhattoa, H. P.; Nissen, P. H.; Orth, M.; Streichert, T.; Young, I. S.; Macintyre, E.; Fraser, A. G.; Neumaier, M., Implementation of the new EU IVD regulation – urgent initiatives are needed to avert impending crisis. Clinical Chemistry and Laboratory Medicine (CCLM) 2022, 60, (1), 33–43.

8. Anastasiou, O. E.; Theodoropoulos, F.; Taube, C.; Fiedler, M.; Dittmer, U., Common respiratory viral infections: Bilateral versus unilateral bronchoalveolar lavage versus endotracheal aspiration. Journal of Medical Virology 2021, 93, (6), 3955–3959.

9. Azadeh, N.; Sakata, K. K.; Saeed, A.; Mullon, J. J.; Grys, T. E.; Limper, A. H.; Binnicker, M. J., Comparison of Respiratory Pathogen Detection in Upper versus Lower Respiratory Tract Samples Using the BioFire FilmArray Respiratory Panel in the Immunocompromised Host. Canadian Respiratory Journal 2018, 2018, 2685723.

10. Azadeh, N.; Sakata Kenneth, K.; Brighton Anjuli, M.; Vikram Holenarasipur, R.; Grys Thomas, E., FilmArray Respiratory Panel Assay: Comparison of Nasopharyngeal Swabs and Bronchoalveolar Lavage Samples. Journal of Clinical Microbiology 2015, 53, (12), 3784–3787.

11. Gao, C. A.; Cuttica, M. J.; Malsin, E. S.; Argento, A. C.; Wunderink, R. G.; Smith, S. B.; Investigators, N. C., Comparing Nasopharyngeal and BAL SARS-CoV-2 Assays in Respiratory Failure. Am J Respir Crit Care Med 2021, 203, (1), 127–129.

12. Geri, P.; Salton, F.; Zuccatosta, L.; Tamburrini, M.; Biolo, M.; Busca, A.; Santagiuliana, M.; Zuccon, U.; Confalonieri, P.; Ruaro, B.; Agaro, P.; Gasparini, S.; Confalonieri, M., Limited role for bronchoalveolar lavage to exclude Covid-19 after negative upper respiratory tract swabs: a multicenter study. European Respiratory Journal 2020, 2001733.

13. Kaku, N.; Hashiguchi, K.; Akamatsu, N.; Wakigawa, F.; Matsuda, J.; Komaru, K.; Nakao, T.; Harada, Y.; Hara, A.; Uno, N.; Sakamoto, K.; Morinaga, Y.; Kitazaki, T.; Hasegawa, H.; Miyazaki, T.; Fukuda, M.; Izumikawa, K.; Mukae, H.; Yanagihara, K., Evaluation of a novel rapid TRC assay for the detection of influenza using nasopharyngeal swabs and gargle samples. European Journal of Clinical Microbiology & Infectious Diseases 2021, 40, (8), 1743–1748.

14. Genelhoud, G.; Adamoski, D.; Spalanzani, R. N.; Bochnia-Bueno, L.; de Oliveira, J. C.; Gradia, D. F.; Bonatto, A. C.; Wassem, R.; Raboni, S. M.; Nogueira, M. B.; de Araujo-Souza, P. S., Comparison of SARS-CoV-2 molecular detection in nasopharyngeal swab, saliva, and gargle samples. Diagnostic Microbiology and Infectious Disease 2022, 103, (2), 115678.

15. Arnold, D.; Suri, T.; Hamilton, F.; Morley, A.; Patole, S.; Medford, A.; Muir, P.; Maskell, N., The role of viruses in the development of pleural infection. European Respiratory Journal 2019, 54, (suppl 63), PA3834.

16. Sevin, C. M.; Peng, S.; Skouras, V.; Lee, W.; Pappas, T.; Gern, J. E.; Light, R. W., Do Viral Infections Cause Pleural Effusions? In C43. PLEURAL DISEASE I -- DIAGNOSTIC ISSUES, American Thoracic Society: 2009; p A4459.

