## Supplement for "Comparison of common respiratory pathogen detection in nasopharyngeal swabs, saline gargle specimens, pleural effusion and lower respiratory tract samples"

|  | Sex | Virus | Ct value | Immunosuppression |
| --- | --- | --- | --- | --- |
| 1 | F | Adenovirus | 35 | - |
| 2 | F | Parainfluenza Virus (Type 1) | 28 | - |
| 3 | M | Adenovirus | 35 | - |
| 4 | M | Bocavirus | 34 | - |
| 5 | M | Rhinovirus | 33 | - |
| 6 | M | Bocavirus | 37 | - |
| 7 | M | Influenzavirus B | 16 | - |
| 8 | M | Adenovirus | 22 | - |
| 9 | M | Rhinovirus | 37 | - |
| 10 | M | Adenovirus | 35 | - |
| 11 | F | Adenovirus | 35 | AML |
| 12 | F | HCoV-OC43 | 34 | AML |
| 13 | F | Bocavirus | 36 | aHSCT |
| 14 | M | Adenovirus | 22 | aHSCT |
| 15 | M | Rhinovirus, Adenovirus | 31 and 32 | Liver transplantation |
| 16 | M | Bocavirus | 26 | ALL |
| 17 | M | Rhinovirus | 27 | Kidney transplantation |
| 18 | M | Rhinovirus | 29 | Chemotherapy (solid tumor) |
| 19 | M | Rhinovirus, HCoV-229E | 31 and 35 | ALL |
| 20 | M | Parainfluenza Virus (Type 3) | 35 | aHSCT |
| 21 | M | Parainfluenza Virus (Type 1) | 31 | Primary immune deficiency |
| 22 | M | Parainfluenza Virus (Type 1) | 33 | Primary immune deficiency |
| 23 | F | Parainfluenza Virus (Type 1) | 28 | Lung transplantation |
| 24 | F | HCoV-229E | 37 | Lung transplantation |
| 25 | M | Rhinovirus | 32 | Lung transplantation |
| 26 | M | Rhinovirus | 24 | Lung transplantation |
| 27 | M | Parainfluenza Virus (Type 4) | 26 | Lung transplantation |
| 28 | M | HCoV-OC43 and Adenovirus | 13 and 31 | Lung transplantation |
| 29 | F | Influenzavirus A | 37 | Lung transplantation |

Table S1: Cases with negative nasopharyngeal swabs (NPS) but positive lower respiratory tract (LTR) samples.

M= male; F=female; HCoV-OC43=human coronavirus OC43; HCoV-229E=human coronavirus 229E; AML=acute myeloid leukemia; ALL= acute lymphoblastic leukemia; aHSCT=allogeneic hematopoietic stem cell transplantation.

Table S2: Cases with discrepant results between corresponding nasopharyngeal swabs (NPS) and saline gargle specimens (SGS).

| id | Sex | Virus | Ct value | Positive Material |
| --- | --- | --- | --- | --- |
| 1 | M | Enterovirus | 35 | NPS |
| 2 | F | HCoV-NL63 | 25 | NPS |
| 3 | F | HMPV | 36 | NPS |
| 4 | M | Rhinovirus | 34 | NPS |
| 5 | M | Rhinovirus | 31 | SGS |
| 6 | F | Bocavirus | 37 | SGS |
| 7 | M | Rhinovirus | 24 | SGS |
| 8 | F | Parainfluenza virus Type 3 | 30 | SGS |
| 9 | M | Rhinovirus and Influenza A | 28 and 34 | SGS |
| 10 | M | Adenovirus | 36 | SGS |

M= male; F=female; HCoV-NL63= human coronavirus NL63; HMPV= human metapneumovirus

Table S3: Cases with discrepant results between corresponding lower respiratory tract (LTR) and pleural effusion (PE) samples.

| id | Sex | Virus | Ct value | Positive Material |
| --- | --- | --- | --- | --- |
| 1 | M | RSV | 21 | BAL |
| 2 | M | Rhinovirus | 16 | EA |
| 3 | M | HCoV-HKU1 | 28 | BAL |
| 4 | F | Adenovirus | 31 | BAL |
| 5 | M | HMPV and Bocavirus | 20 and 27 | BAL |
| 6 | M | Influenza B | 35 | PE |

M= male; F=female; HCoV-HKU1= human coronavirus HKU1; HMPV= human metapneumovirus; BAL=bronchioalveolar lavage; EA= endotracheal aspirate; PE=pleural effusion.
